# Predicting the number of people infected with SARS-COV-2 in a population using statistical models based on wastewater viral load

**DOI:** 10.1101/2020.07.02.20144865

**Authors:** Juan A. Vallejo, Soraya Rumbo-Feal, Kelly Conde-Pérez, Ángel López-Oriona, Javier Tarrío-Saavedra, Rubén Reif, Susana Ladra, Bruno K. Rodiño-Janeiro, Mohammed Nasser, Ángeles Cid, María C Veiga, Antón Acevedo, Carlos Lamora, Germán Bou, Ricardo Cao, Margarita Poza

## Abstract

The quantification of the SARS-CoV-2 RNA load in wastewater has emerged as a useful tool to monitor COVID-19 outbreaks in the community. This approach was implemented in the metropolitan area of A Coruña (NW Spain), where wastewater from a treatment plant was analyzed to track the epidemic dynamics in a population of 369,098 inhabitants. Statistical regression models from the viral load detected in the wastewater and the epidemiological data from A Coruña health system that allowed us to estimate the number of infected people, including symptomatic and asymptomatic individuals, with reliability close to 90%, were developed. These models can help to understand the real magnitude of the epidemic in a population at any given time and can be used as an effective early warning tool for predicting outbreaks. The methodology of the present work could be used to develop a similar wastewater-based epidemiological model to track the evolution of the COVID-19 epidemic anywhere in the world.

## 1. INTRODUCTION

According to previous reports, a great proportion of patients infected with SARS-CoV-2 are asymptomatic (Bi et al. 2020, Day 2020, Randazzo et al. 2020a, Yang et al. 2020), a condition that depends on many factors such as the mean age in the population and that promotes the undetected spread of COVID-19. A systematic literature review found that at least an important proportion of COVID-19 patients, including symptomatic and asymptomatic people, tested for fecal viral RNA were positive from initial steps of infection (Gupta et al, 2020). This excreta of viral RNA in patients stool occurs for an extended period, importantly even more than a month after the patient has tested negative for their respiratory samples (Chen et al. 2020, Wölfel et al. 2020, Wu et al. 2020a, Xing et al. 2020, Xu et al. 2020, Zhang et al. 2020). Therefore, genetic material of SARS-CoV-2 can be found in wastewater (Lodder and de Roda Husman 2020), which has made monitoring of viral RNA load in sewage an excellent tool for the epidemiological tracking of the actual pandemic as well as an extremely efficient early warning tool for outbreaks detection (Randazzo et al. 2020a, Ahmed et al. 2020, Medema et al. 2020, Peccia et al. 2020, Wu et al. 2020b, Wurtzer et al. 2020).

Wastewater is a dynamic system and its analysis can provide a faithful reflection of the circulation of microorganisms in the population. Previous studies have evaluated the presence in wastewater of other viruses, such as enterovirus, norovirus, hepatitis A virus, adenovirus, poliovirus or sapovirus (Ehlers et al. 2005, Hellmér et al. 2014, Hovi et al. 2012, Lizasoain et al. 2018, Mancini et al. 2019). During the present global COVID-19 pandemic, processes to monitor SARS-CoV-2 in wastewater were first developed in the Netherlands (Medema et al. 2020), followed by the USA (Nemudryi et al. 2020), France (Wurtzer et al. 2020), Australia (Ahmed et al. 2020), Italy (La Rosa et al. 2020) and Spain (Randazzo et al. 2020a, Randazzo et al. 2020b). In the first study to look at SARS-CoV-2 in wastewater, seven cities and Schiphol airport in the Netherlands were monitored during the early stages of the pandemic. Genetic material started to appear at more sites over time, as the number of cases of COVID-19 increased (Medema et al. 2020). In the USA, a wastewater plant in Massachusetts detected a viral RNA load higher than expected based on the number of confirmed cases, reflecting viral shedding of asymptomatic cases in the community (Wu et al. 2020b). Three wastewater plants in Paris measured the concentration of the virus over a 7-week period that included the beginning of the lockdown on March 17^th^. They found that viral RNA load was correlated with the number of confirmed COVID-19 cases. They also noted that viral RNA could be detected in the wastewater before the exponential growth of the disease and that the amount of viral RNA decreased as the number of COVID-19 cases went down, roughly following an eight-day delay (Wurtzer et al. 2020). Another study of six wastewater plants in Spain, covering a region with the lowest prevalence of COVID-19, also detected the virus in wastewater before the first COVID-19 cases were reported (Randazzo et al. 2020a). A recent study from Yale University took a different approach by measuring the concentration of SARS-CoV-2 RNA in sewage sludge in New Haven (Connecticut, USA), as opposed to wastewater. They tracked viral RNA concentrations against hospital daily admissions and confirmed COVID-19 cases in the community. They found that viral RNA concentrations were highest 3 days before peak hospital admissions, and 7 days before peak community COVID-19 cases, which showed that virus RNA concentration is an earlier indicator of progression of COVID-19 in the community than traditional epidemiological indicators (Peccia et al. 2020). Also, the presence of SARS-CoV-2 in sludge at concentrations potentially suitable for monitoring was confirmed (Balboa et al. 2020). These studies showed the potential of monitoring SARS-CoV-2 levels in wastewater and sewage sludge to track and even pre-empt outbreaks in the community.

During the last decade, Wastewater-Based Epidemiology (WBE) has emerged as a highly relevant discipline with the potential to provide objective information by combining the use of cutting-edge analytical methodologies with the development of *ad hoc* modelling approaches. WBE has been extensively used to predict with high accuracy the consumption patterns of numerous substances, such as the use of illicit drugs in different populations or countries (EMCDDA 2020). Therefore, the development of epidemiology models based on wastewater analysis has been intensive in the last years. Several examples from the literature showed different approaches and strategies to tackle the uncertainty associated with WBE studies. For example, Goulding and Hickman assumed three main sources of uncertainty (fluctuations in flow, uncertainty in analytical determinations, and the actual size of the population served by the wastewater treatment plant) and, using Bayesian statistics, fitted the data to linear regression hierarchical models (Goulding and Hickman 2020). Other modelling approaches (Croft et al. 2020) considered Monte Carlo simulations to deal with uncertainties and with the propagation of errors associated with the parameters that are usually considered in WBE. Again, wastewater inflow variability was highlighted as a prominent source of uncertainty, as well as the stability of the substances in wastewater and their pharmacokinetics. In general, WBE studies showed that despite the wide number of parameters involved in predicting the consumption rate of a specific substance, a correct selection of assumptions combined with a thoughtful modelling process will overcome such uncertainty, leading to accurate results. Recently, a study has confirmed the theoretical feasibility of combining WBE approaches with SARS-CoV-2/COVID-19 data (Hart and Halden 2020).

The main objective of the present work was to develop a useful statistical model to determine the entire SARS-CoV-2 infected population, including symptomatic and asymptomatic people, as well as to predict outbreaks, by tracking the viral load present in the wastewater of a treatment plant located in the Northwest of Spain that serves a metropolitan area with near 370,000 residents.

## 2. MATERIAL AND METHODS

### 2.1. Sample Collection

The wastewater treatment plant (WWTP) Bens (43° 22’ 8.4’’ N 8° 27’ 10.7’’ W, A Coruña, Spain) serves the metropolitan area of A Coruña, which includes the municipalities of A Coruña, Oleiros, Culleredo, Cambre and Arteixo, which correspond to a geographical area of 277.8 km^2^ and to a population of 369,098 inhabitants (Figure 1). The wastewater samples were collected by automatic samplers installed both at the entrance of the WWTP Bens and in a sewer collecting sewage from COVID-19 patients housed on 7 floors of the University Hospital of A Coruña (CHUAC). At the WWTP Bens, 24-h composite samples were collected from April 15^th^ until June 4^th^, while at CHUAC 24-h composite samples were collected from April 22^nd^ to May 14th (Dataset S1). In addition, samples were collected at 2-h intervals for 24 h on specific days at the WWTP Bens and at CHUAC (Dataset S1). The 24-h composite samples were collected by automatic samplers taking wastewater every 15 min in 24 bottles (1 h per bottle) and, when the 24-h collection ended, the 24 bottles were integrated and a representative sample of 100 mL was collected. For the collection of 2 h intervals, 2 bottles obtained every 2 h were integrated, finally providing 12 bottles per day.

**Figure 1.**
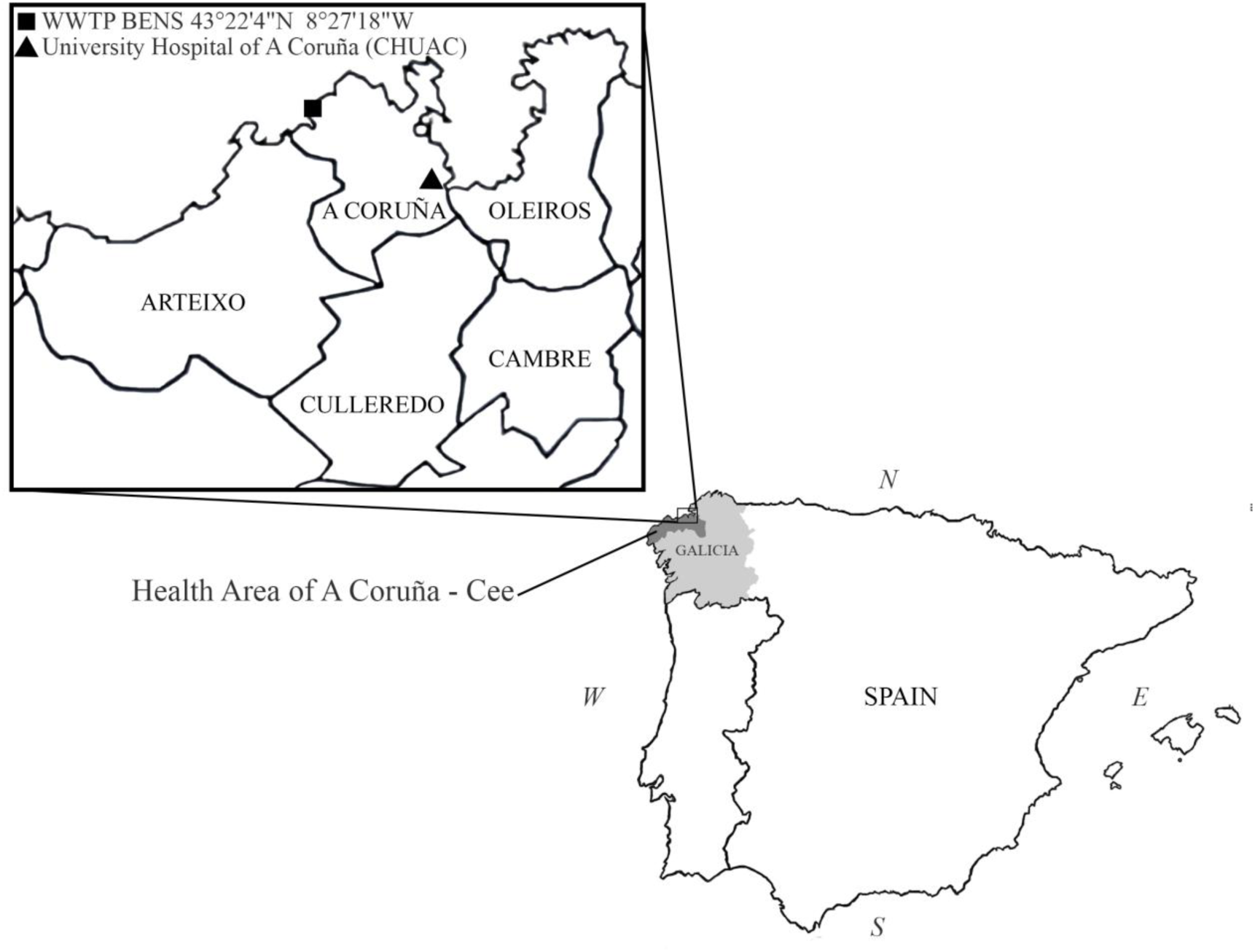
Map showing the Galician region in Spain, the metropolitan area of A Coruña including Oleiros, Cambre, Culleredo, Arteixo and A Coruña municipalities, and the Health Area of A Coruña-Cee, as well as the specific locations of the University Hospital of A Coruña (CHUAC) and WWTP Bens.

### 2.2. Sample processing

Samples of 100 mL were processed immediately after collection at 4 °C. Firstly, 100 mL samples were centrifuged for 30 min at 4000 *x g* and then filtered through 0.22 µm membranes. Samples were then concentrated and dialyzed using Amicon Ultra Filters 30 KDa (Merck Millipore) in 500 µL of a buffer containing 50 mM Tris-HCL, 100 mM NaCl y 8 mM MgSO_4_. Samples were preserved in RNAlater reagent (Sigma-Aldrich) at −80 °C.

### 2.3. RNA extraction and qRT-PCR assays

RNA was extracted from the concentrates using the QIAamp Viral RNA Mini Kit (Qiagen, Germany) according to manufacturer’s instructions. Briefly, the sample was lysed under highly denaturing conditions to inactivate RNases and to ensure isolation of intact viral RNA. Then, the sample was loaded in the QIAamp Mini spin column where RNA was retained in the QIAamp membrane. Samples were washed twice using washing buffers. Finally, RNA was eluted in an RNase-free buffer. The quality and quantity of the RNA was checked using a Nanodrop Instrument and an Agilent Bioanalyzer. Samples were kept at −80°C until use.

RT-qPCR assays were done in a CFX 96 System (BioRad, USA) using the qCOVID-19 kit (GENOMICA, Spain) through N gene (coding for nucleocapsid protein N) amplification. Reaction mix (15 µL) consisted of: 5 µL 4x RT-PCR Mix containing DNA polymerase, dNTPs, PCR buffer and a VIC internal control; 1 µL of Reaction Mix 1 containing primers and FAM probe for N gene; and 0.2 µL of reverse transcriptase enzyme. The internal control allowed discarding the presence of inhibitors. The cycling parameters were 50 °C for 20 minutes for the retrotranscription step, followed a PCR program consisting of a preheating cycle of 95 °C for 2 min, 50 cycles of amplification at 95 °C for 5 s and finally one cycle of 60 °C for 30 s. RT-qPCR assays were done in sextuplicate.

For RNA quantification, a reference pattern was standardized using the Human 2019-nCoV RNA standard from European Virus Archive Glogal (EVAg) (Figure S1). To build the calibration curve, the decimal logarithm of SARS-CoV-2 RNA copies/µL ranging from 5 to 500 were plotted against Ct (threshold cycle) values. Calibration was done amplifying the N gene.

### 2.4. Data collection

The model was fully customized to the scenario of the metropolitan area of A Coruña integrating data gathered from different sources:

Daily observations at the meteorological station of A Coruña-Bens for the period March 1^st^ – May 31^st^, 2020, including rainfall, temperature, and humidity (source: Galician Meteorology Agency, MeteoGalicia (Dataset S2)).

Cumulative and active number of COVID-19 cases in the metropolitan area of A Coruña and from the health area A Coruña – Cee for the period March 1^st^ – May 31^st^ (source:Galician Health Service (SERGAS), the General Directorate of Public Health (Automous Government of Galicia) and the University Hospital of A Coruña (CHUAC) (Dataset S3).) Since flow may be an important variable when determining the viral load in the wastewater, an exploratory data analysis for the volume of water pumped at the WWTP Bens during the lockdown period has been performed using flow data (Dataset S4). This flow study is described in the Supplementary Material section.

### 2.5. Backcasting of COVID-19 active cases

Preliminary statistical methods have been devised to backcast the number of COVID-19 active cases based on reported official cases.

Follow-up times (available only until May 7^th^) for anonymized individual reported COVID-19 cases in Galicia (NW Spain where the WWTP Bens is located, Figure 1) have been used to count the number of cases by municipality based on patient zip codes. Since the epidemiological discharge time is missing, the number of active cases in the metropolitan area of A Coruña could not be obtained but the cumulative number of cases was computed. On the other hand, the main epidemiological series for COVID-19 were publicly available in Galicia at the level of health areas. However, the definition of one of the series changed from cumulative cases to active cases in April 29^th^.

Thus, the epidemiological series for COVID-19 in the health area of A Coruña – Cee (population 551,937) was used to estimate the epidemiological series for COVID-19 for the metropolitan area of A Coruña (population 369,098). To do this, a linear regression model was used to relate the relative cumulative and active cases (cases per million) of COVID-19 for the health area of A Coruña – Cee. Predicting the rate of active cases and considering the population size in the metropolitan area gives the estimated total number of official active cases in the five municipalities.

The previous approach is only possible until May 7^th^, our database update date. To estimate the number of official active cases from May 8^th^ onwards, another linear regression model has been used to relate the number of active cases in the health area of A Coruña – Cee and in the metropolitan area of A Coruña. Since the number of active cases in the health area has been reported until June 5^th^, the series of estimated official active cases could be backcasted from May 8^th^ until June 5^th^.

Finally, to transform the official number of COVID-19 cases into the real number, the ratio mean of real cases / mean of official cases was estimated using the official figures of cumulative cases. The results of the seroprevalence study carried out by the National Center of Epidemiology in Spain were used to estimate the number of actual active cases in Galicia: 56,713 for April 27^th^ – May 11^th^ (prevalence 2.1%) and 59,414 for May 18^th^ – June 1^st^ (prevalence 2.2%) (Pollán et al. 2020). Confronting these numbers with the official numbers in May 11^th^ (10,669) and June 1^st^ (11,308) gives estimated ratios of 5.316 and 5.254 in these two periods, with an average of around 5.29. This conversion factor was used to backcast the series of real active cases based on the estimated daily official COVID-19 cases in the metropolitan area of A Coruña. Some of these series, including the backcasted series of real active cases, are included in the Dataset S3.

### 2.6. Nonparametric setting of viral load overtime

Generalized Additive Models (GAM) using a basis of cubic regression splines (Hastie et al. 1990) and LOESS (Cleveland 1979) nonparametric regression models have been used to fit the viral load along the day on May 5^th^, 6^th^, 11^th^ and 12^th^, and as a function of time at CHUAC from April 22^nd^ to May 12^th^ and at WWTP Bens from April 16^th^ to June 3^rd^. Several outliers have been removed from the data, corresponding to unexpected and intensive pipeline cleaning episodes (8-hour 70 °C water cleaning during Thursday-Friday nights) carried out in April 23^rd^-24^th^, April 30^th^ - May 1^st^ and May 7^th^-8^th^.

### 2.7. Viral load models

In order to find a useful statistical model to predict the number of real infected cases, including symptomatic and asymptomatic people, well-known regression models, such as simple and multivariate linear models, and more flexible models, such as nonparametric (e.g. local linear polynomial regression) and semiparametric (GAM and LOESS) models, have been formulated. The flexible ones allowed the introduction of linear and smooth effects of the predictors on the response.

All these models have been successfully used to predict the number of COVID-19 active cases based on the measured viral load (number of RNA copies/L) at WWTP Bens, daily flow in the sewage network as well as other environmental variables, such as rainfall, temperature and humidity.

Diagnostic tests (Q-Q plots, residuals versus fitted values plots and Cook’s distance) were used for outlier detection, which improved the models fit. The R statistical software was used to perform statistical analyses (R Core Team, available at https://www.R-project.org/). Namely, the mgcv library (Wood 2006) was applied to fit GAM models and ggplot2 and GGally (Schloerke et al. 2020, Wickham 2016) to perform correlation analysis, obtain graphical output and fit LOESS models, respectively. The caret R package was used to fit and evaluate regression models.

Although some RT-qPCR replicates could not be measured when the viral load was scarce, due to the limitation of the detection technique (errors randomly occur when the number of copies/L is under 10,000), 74% of the assays led to three or more measured replications, which gives a good statistical approach. However, conditional mean imputation (Enders 2010) was used for unmeasured replications. Thus, unmeasured replications in an assay were replaced by the sample mean of observed measurements in that assay. In the only assay with all (six) unmeasured replications, the number of RNA copies was imputed using the minimum of measured viral load along the whole set of assays.

## 3. RESULTS

### 3.1. Estimated COVID-19 positive cases in the metropolitan area of A Coruña

To model the viral load, the number of COVID-19 positive cases needed to be reported or estimated (Figure 2). However, due to difficulties in determining the exact number of positive cases, mathematical models had to be developed based on data recovered in Dataset S3. Thus, linear regression models (Figure 3) were successfully used to estimate COVID-19 positive cases in the A Coruña – Cee health area, where the region of this study is located (Figure 1). This was based on Intensive Care Unit (ICU) patients before April 29^th^ (Figure 3A) and, from this date on, on positive cases reported by health authorities in Galicia. A linear regression trend was fitted to predict the proportion of positive cases in the health area of A Coruña – Cee based on the proportion of cumulative cases (Figure 3B). This linear regression fit was finally used to estimate the positive cases in the metropolitan area of A Coruña served by the WWTP Bens, by means of the proportion of cumulative positive cases in the same area, which was directly obtained from the individual patient data.

**Figure 2.**
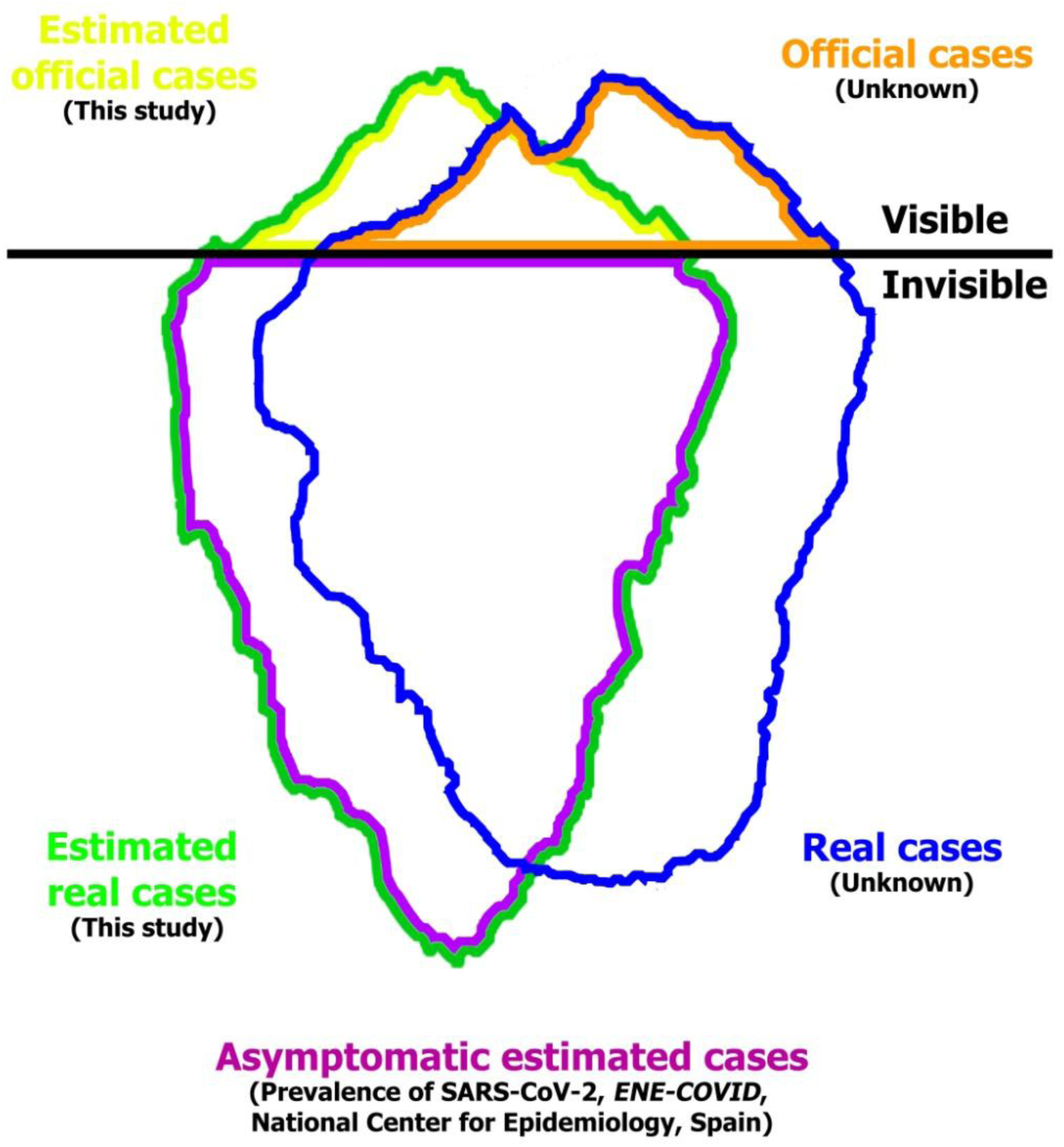
Iceberg representing the overall health of the population of the metropolitan area of A Coruña infected by SARS-CoV-2, showing the real and official cases estimated in this study.

**Figure 3.**
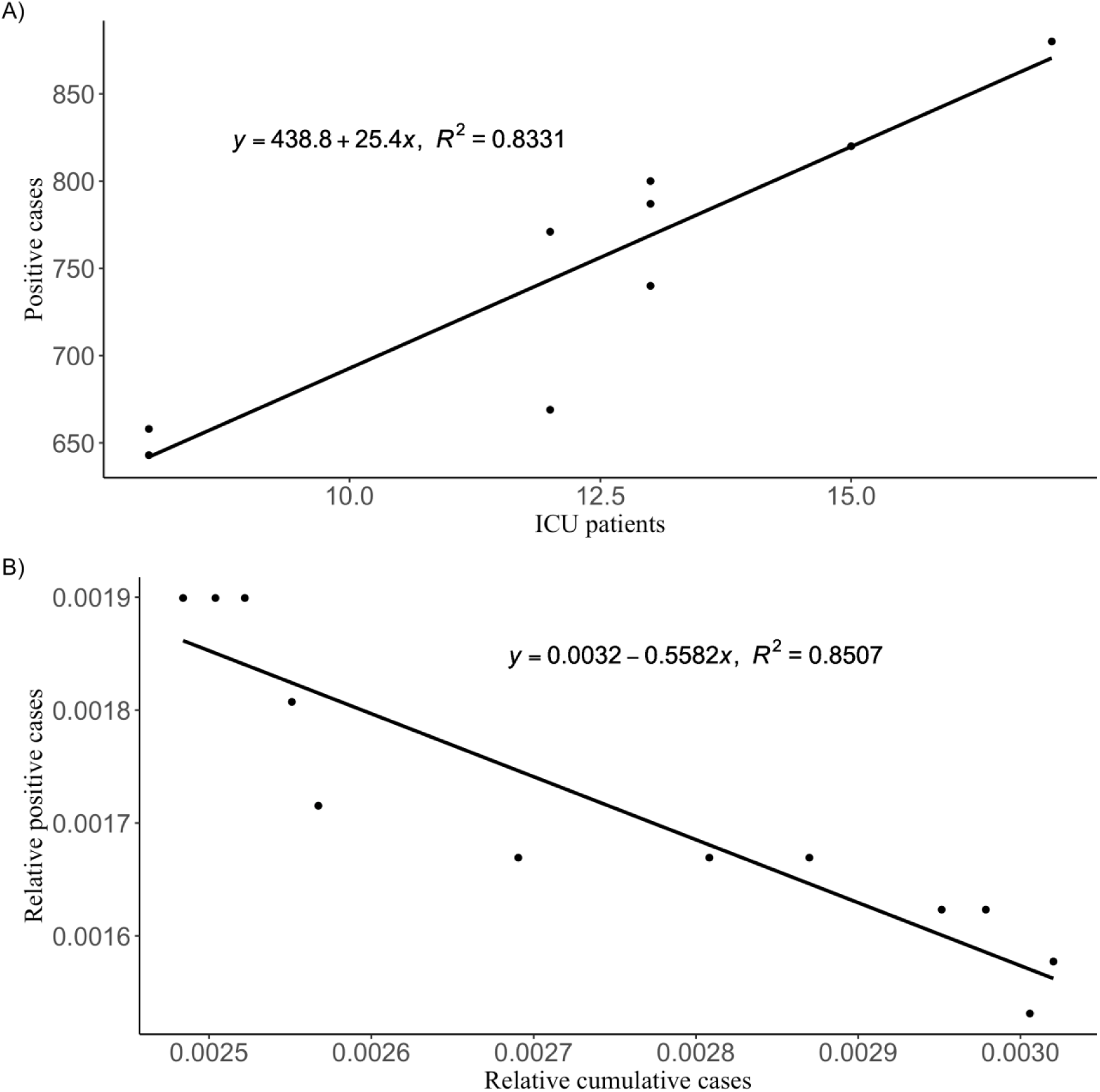
Estimation of the COVID-19 positive cases using simple linear regression models. A) ICU patients versus COVID-19 positive cases in A Coruña – Cee health area, and its linear fit for the period April 29^th^ on. B) Relative cumulative cases versus relative positive cases in the health area of A Coruña – Cee and their linear fit.

### 3.2. Daily variation of SARS-CoV-2 RNA load in the metropolitan area of A Coruña and hospital

The evolution of the viral load along the day both at the A Coruña metropolitan area and at the University Hospital Complex of A Coruña (CHUAC) was monitored in order to discern which amount of the viral load detected in the WWTP Bens came from the CHUAC and which from the community. An analysis of the viral load of the 24-h and 2-h samples collected at the WWTP Bens and CHUAC, as reflected in Dataset S1, was performed. The RT-qPCR results for 24-h samples are included in Dataset S5 and results for 2-h samples are included in Dataset S6. Because of the small sample size, nonparametric LOESS models were used in order to prevent the possible overfitting of alternatives such as GAM. Figure 4 shows the viral load trends at CHUAC (Figure 4A) and at WWTP Bens (Figure 4B) depending on the hour of the day, during four different days. Figure 4A shows the hourly trend at CHUAC, with a maximum around 08:00, whereas the viral load curves at WWTP Bens (Figure 4B) attained a minimum around 05:00 and a maximum between 14:00 and 15:00.

**Figure 4.**
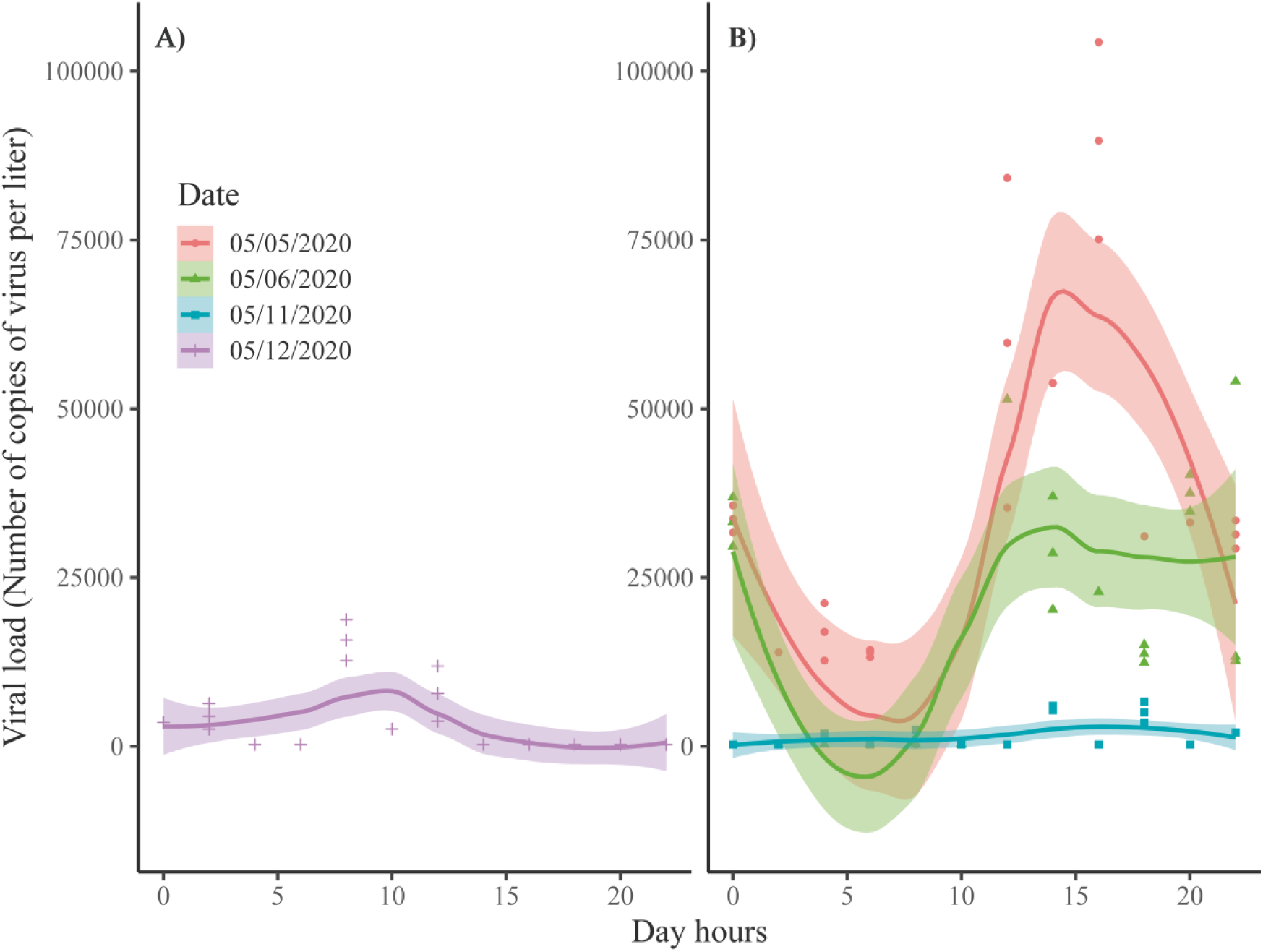
Viral load trend during the day at CHUAC and Bens. A) Viral load with respect to the hour of the day in CHUAC during the 05/12/2020 and nonparametric LOESS fitted model (span parameter equal to 0.75) with 95% confidence interval. B) Viral load with respect to the hour of the day in Bens for three different days in May, and nonparametric models (span parameter equal to 0.75) with 95% confidence interval fitted to the data of each day separately.

### 3.3. Lockdown de-escalation in the metropolitan area of A Coruña

As expected, the mean viral load decreased with time when measured at CHUAC (Figure 5A, late April – mid May) and at WWTP Bens (Figure 5B, mid April – early June) following an asymptotic type trend (fitted using GAM with cubic regression splines).

**Figure 5.**
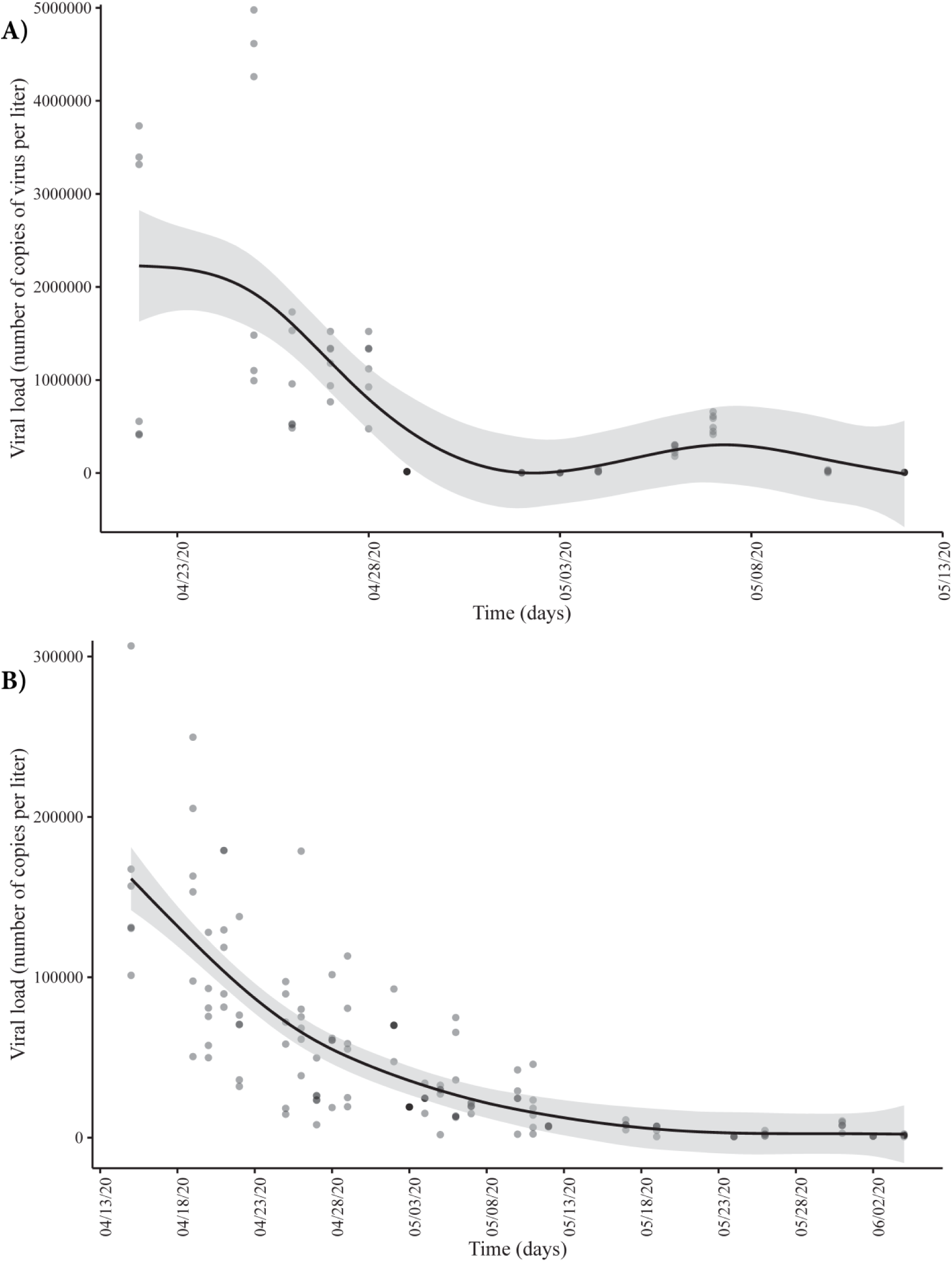
Date effect in the viral load using a nonparametric estimator (GAM) A) at CHUAC and B) at WWTP Bens.

Time course quantitative detection of SARS-CoV-2 in A Coruña WWTP Bens wastewater correlated with the estimated number of COVID-19 positive cases, as shown in Figure 6. The number of copies of viral RNA per liter decreased from around 500,000 to less than 1,000, while the estimated cases of patients infected by SARS-CoV-2 decreased approximately 6-fold in the same period, reaching in both cases the lowest levels in the metropolitan area at the beginning of June.

**Figure 6.**
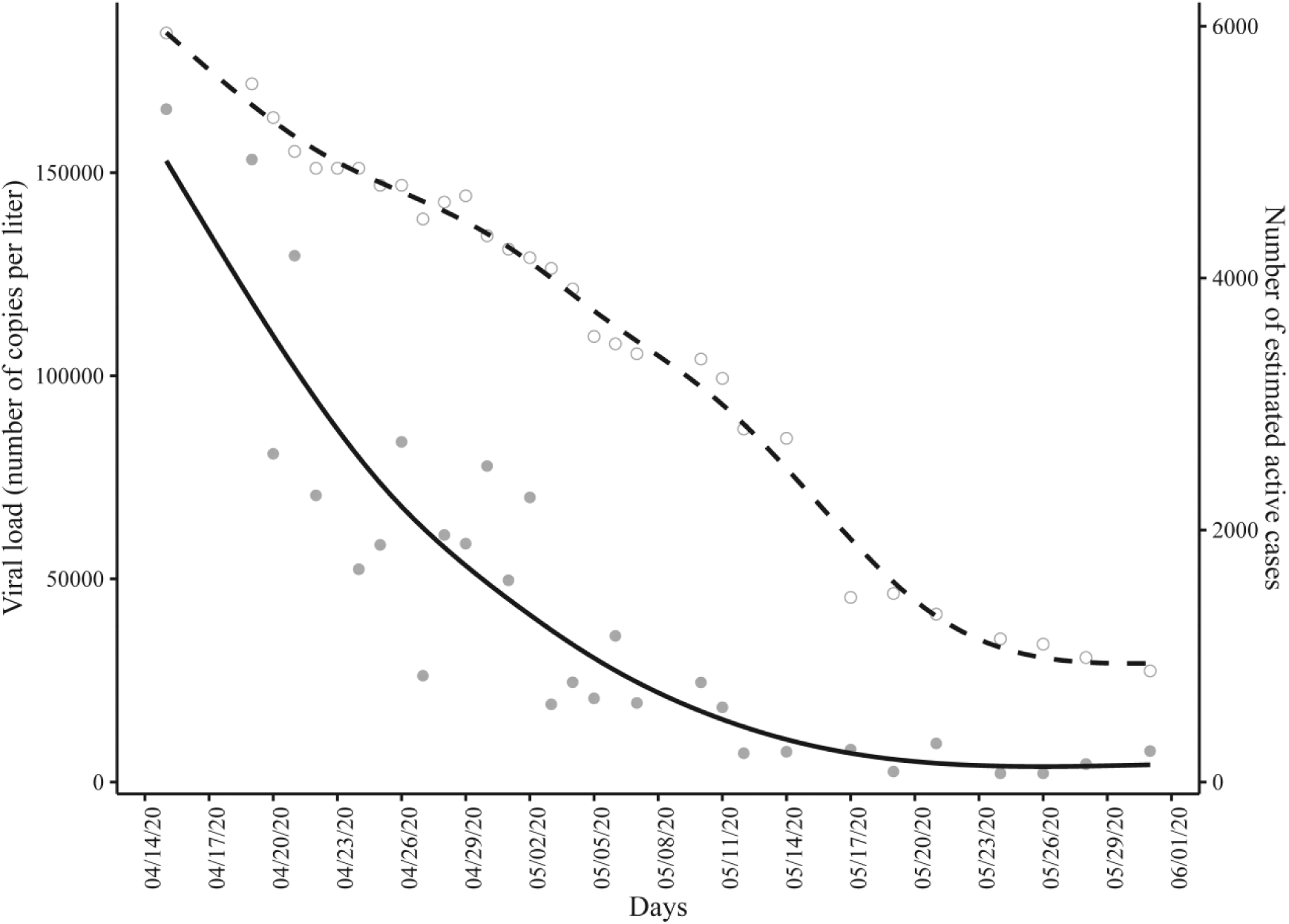
Viral load detected in the influent of the WWTP Bens (solid line) and the estimated number of COVID-19 positive cases (dashed line) in the metropolitan area of A Coruña.

### 3.4. Wastewater epidemiological models based on viral load for COVID-19 real active cases prediction

Firstly, a correlation analysis (Figure 7) was done finding that estimated number of COVID-19 positive cases strongly correlated linearly with the logarithm of daily mean viral load at Bens (R=0.923) and with the mean flow (R=-0.362). Nonetheless, a strong inverse linear relationship between estimated COVID-19 positive cases and time was found (R=-0.99).

**Figure 7.**
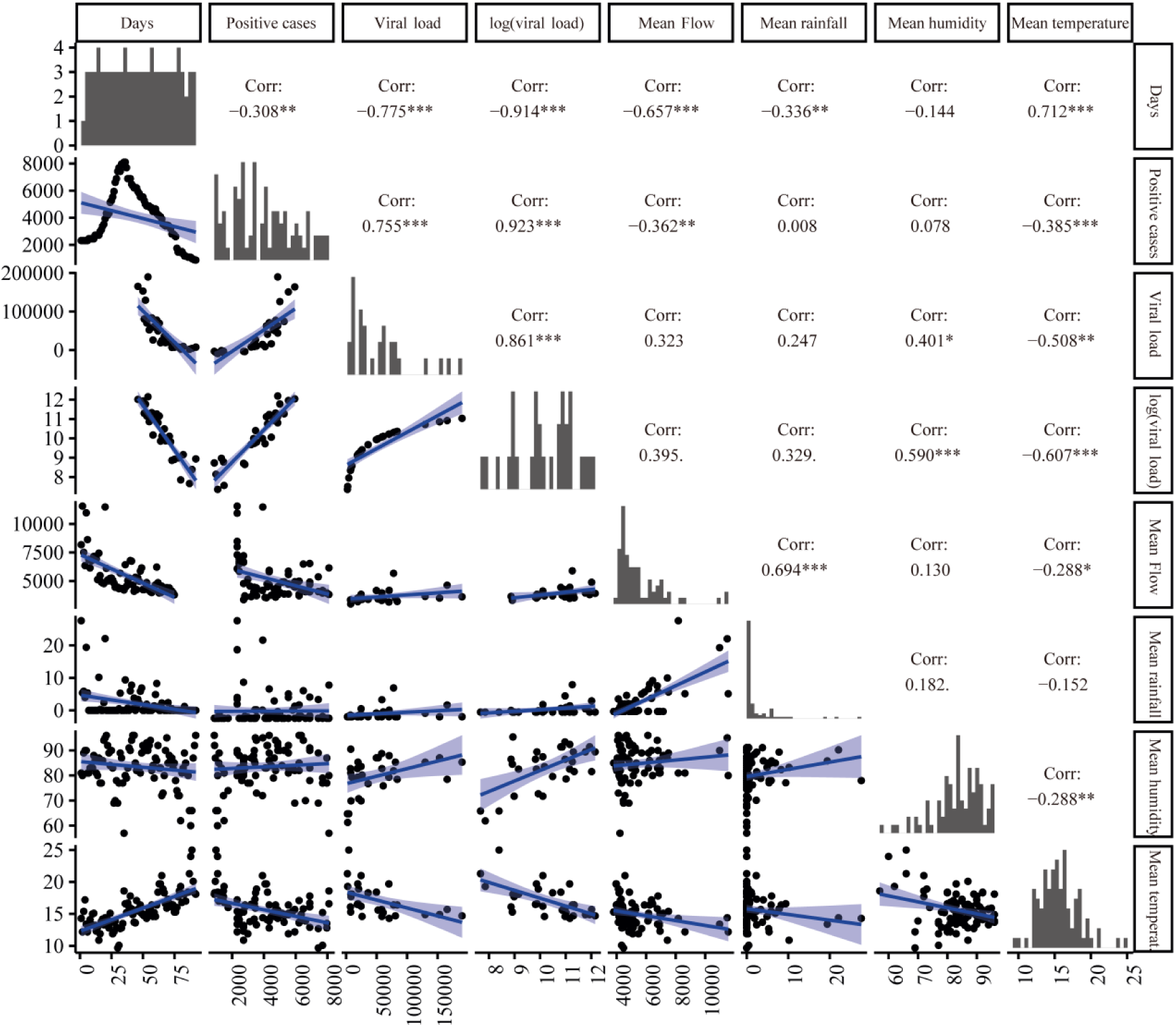
Correlation analysis between estimated COVID-19 positive cases and different variables. Scatterplot matrix shows fitted linear models and linear correlation coefficients of each pair of variables: time (measured in days from the beginning of reported COVID-19 cases), estimated COVID-19 positive cases (positive cases), daily mean viral load measured in Bens, mean flow of sewage water in Bens, mean rainfall, daily mean humidity and daily mean temperature.

Then, different regression models were tested to predict the real number of COVID-19 active cases based on the viral load and the most relevant atmospheric variables. The best results were obtained using GAM models depending on the viral load and the mean flow (Figure 8). The effect of the viral load on the COVID-19 estimated active cases showed a logarithmic shape (Figure 8A), which suggested that the number of COVID-19 real active cases could be modeled linearly as a function of the logarithm of the viral load. On the other hand, the shape of the effect of the mean flow on the estimated number of COVID-19 real active cases appears to be quadratic (Figure 8B), but its confidence band was wide and contained the horizontal line with height zero, which means that the effect of the mean flow was not significant (*p* value=0.142). Therefore, the only independent variable that was significant was the viral load, with R^2^=0.86.

**Figure 8.**
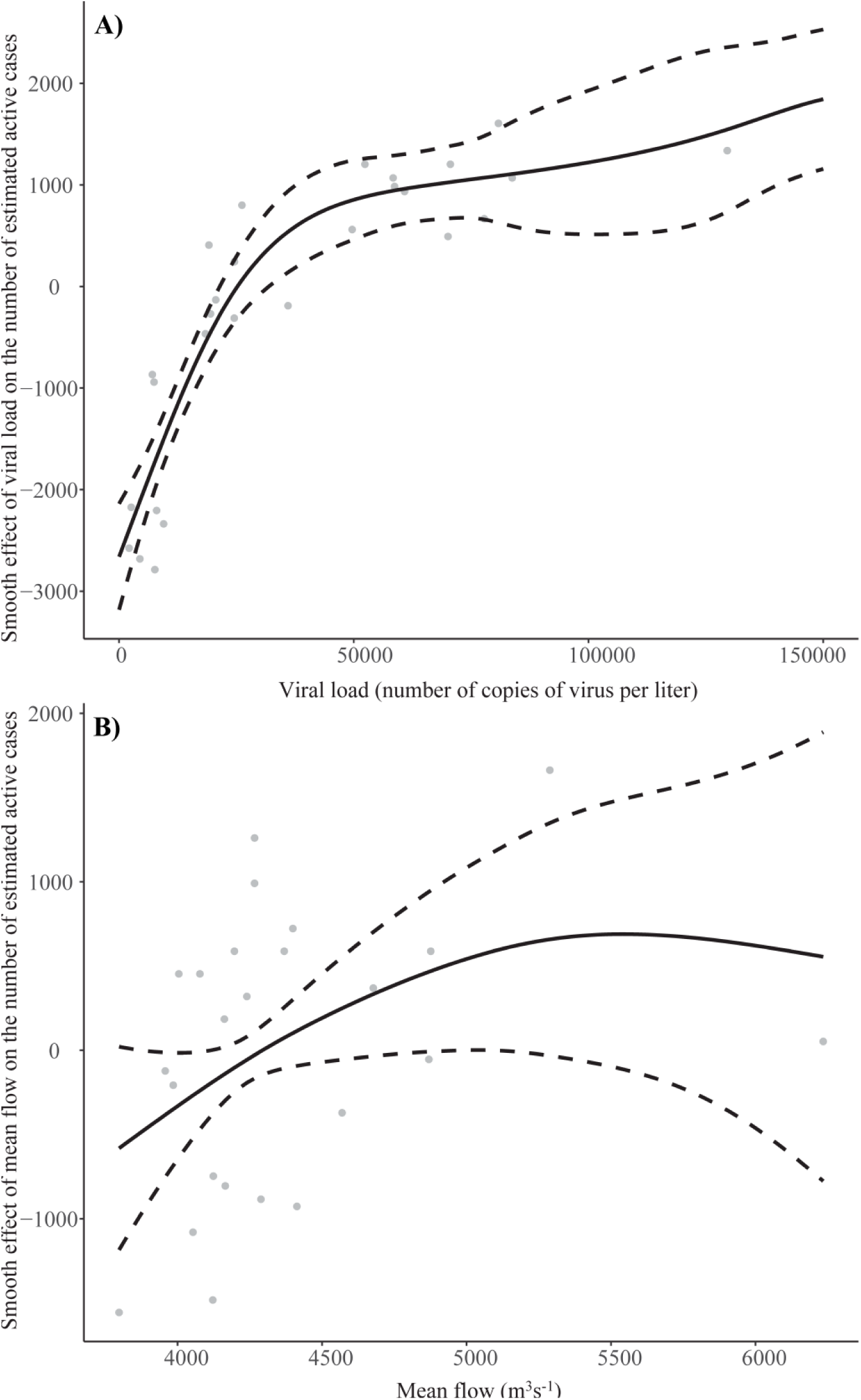
Smooth effect (black line) and confidence band (black stripes) for the viral load (A) and for the mean flow (B) on the number of estimated COVID-19 active cases when fitting a GAM.

Since the nonparametric estimation of the viral load effect had a logarithmic shape, a multiple linear model was fitted using the logarithmic transformation of the viral load, daily flow, rainfall, temperature, and humidity. Figure 9 shows the more explicative models for different number of predictors using the R^2^ maximization criterion, finding that the only significant predictor was the viral load. In fact, when a multivariate linear model depending on three predictors (viral load, daily flow, and rainfall) was fitted, data showed that the only significant explanatory variable was the viral load (*p* value=1.32·10^−8^). Table 1 shows that the effect of the other two predictors, daily flow (*p* value=0.186525) and rainfall (*p* value=0.099239), were not clearly significant.

**Table 1.**
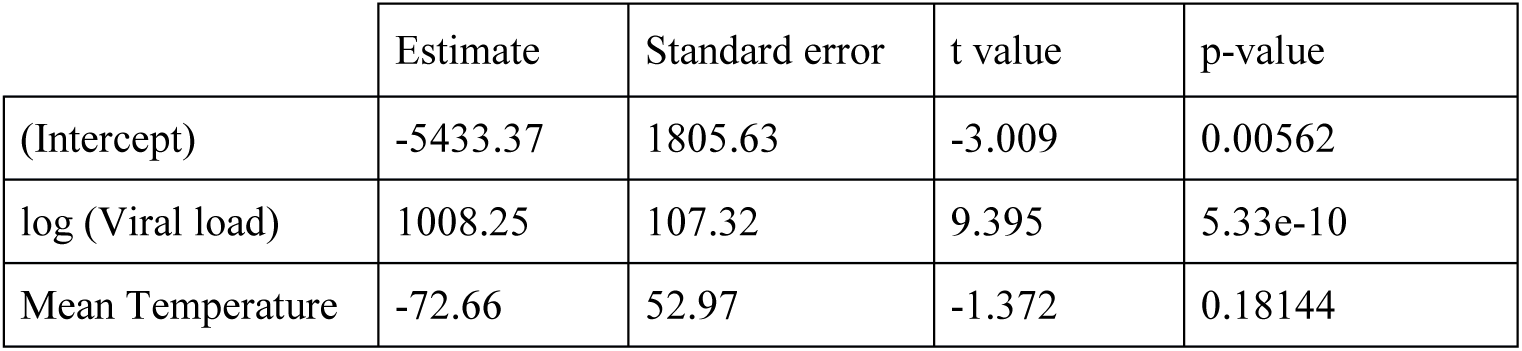
Signification analysis of the multivariate linear model to explain the number of active cases as a function of viral load, and mean temperature.

**Figure 9.**
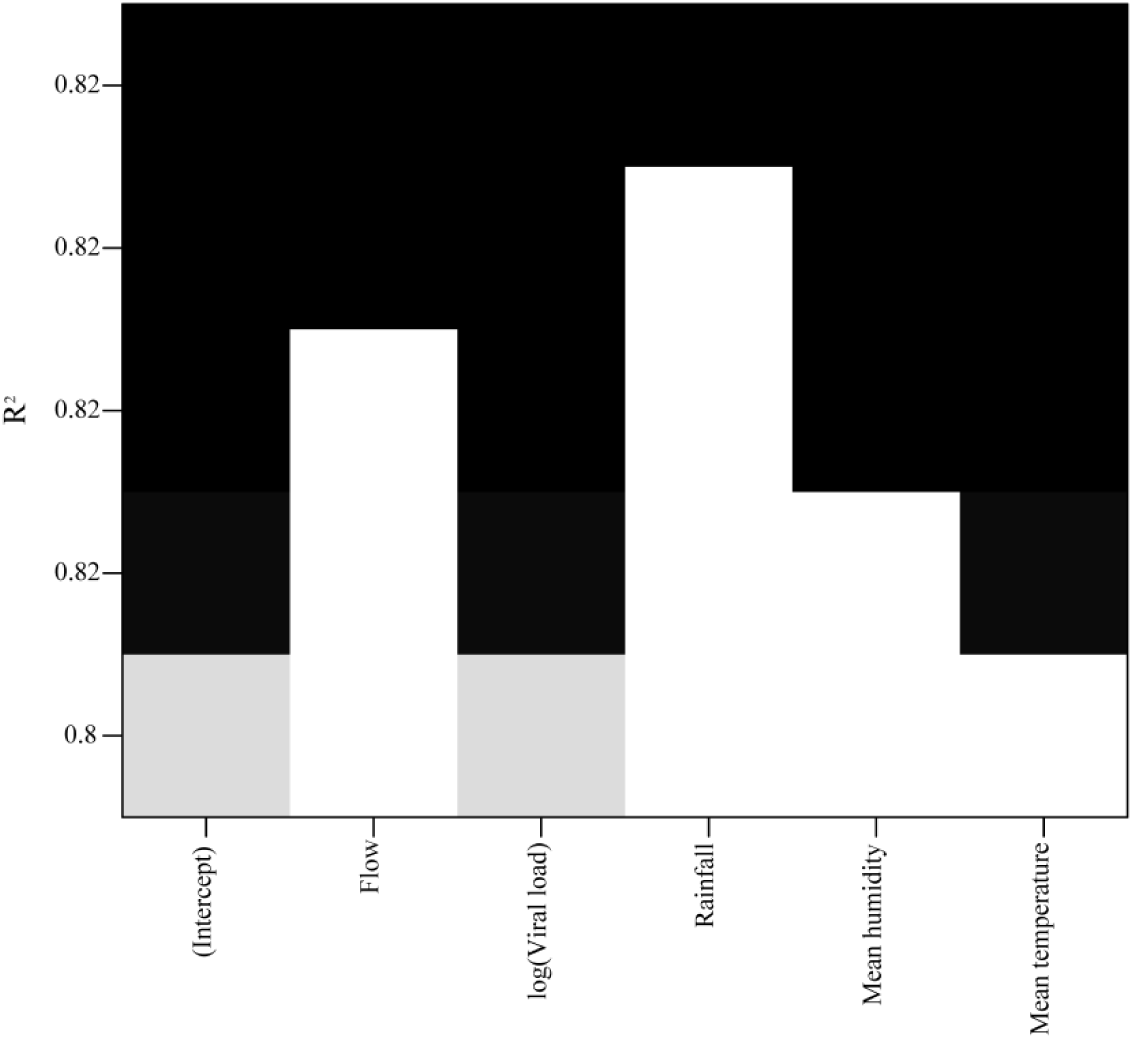
Multivariate linear model selection using the R^2^ maximization criterion. Each row corresponds with the best model using from one to five predictors. The color of the row is darker for higher values of R^2^.

Finally, ignoring the rest of the explanatory variables, only the natural logarithm of the viral load gave a good linear model fit (R^2^=0.851) that was useful to predict the real number of active COVID-19 cases (Figure 10A). After removing three outliers, the fit improved slightly (R^2^=0.894), as shown in Figure 10B.

**Figure 10.**
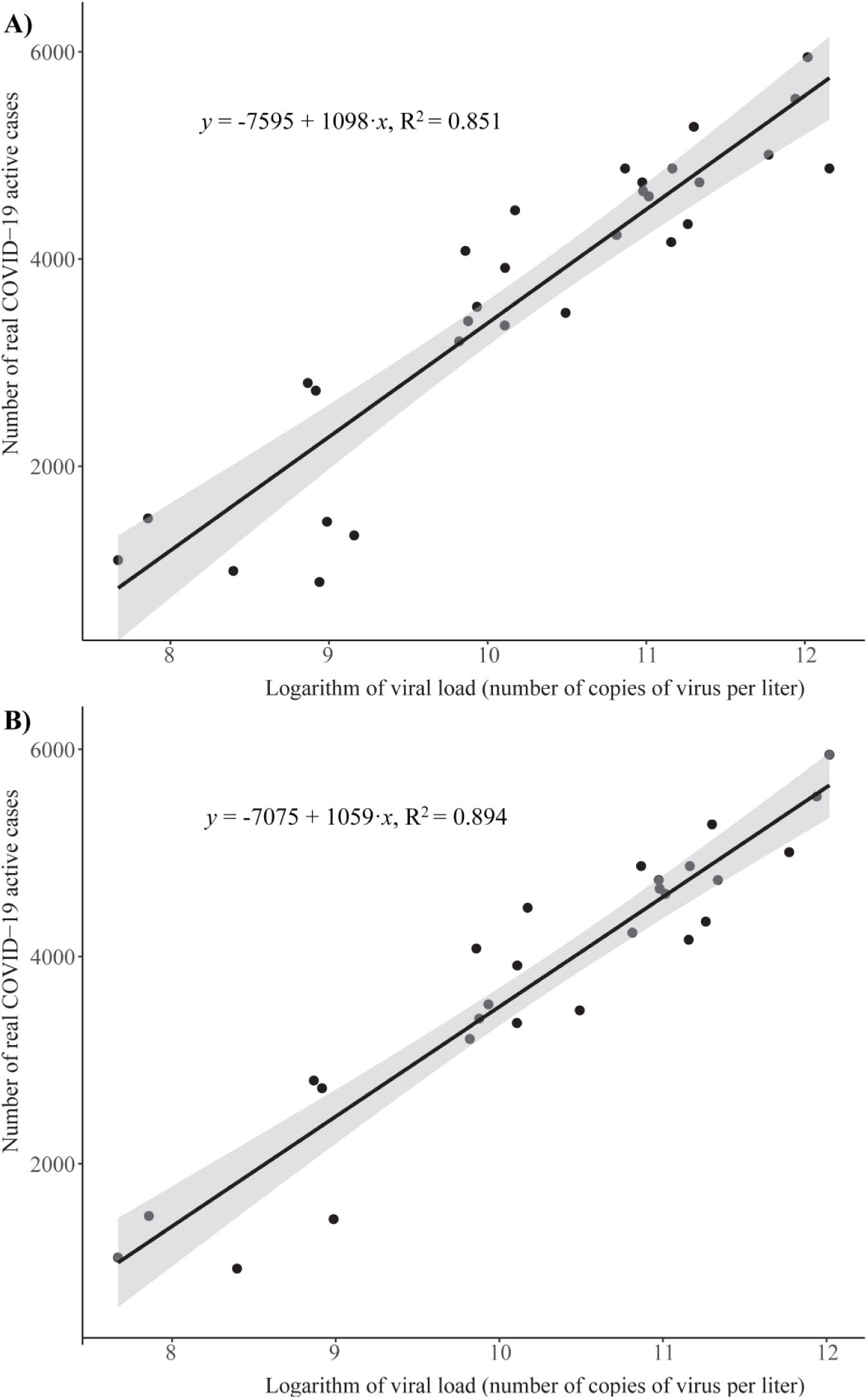
Estimation of the number of COVID-19 real active cases using a linear regression model. Scatterplot represents the logarithm of the viral load measured in WWTP Bens and the estimated number of COVID-19 real active cases before (A) and after (B) removing the three outliers detected. The linear fit (black line) and the confidence band (grey shaded area) are also included.

The final fitted linear model became:

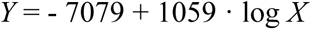

where *Y* denotes the real number of active COVID-19 cases, *X* is the viral load (number of RNA copies per L) and log stands for the natural logarithm.

For instance, a viral load of *Y* = 150,000 copies per liter would lead to an estimated number of *X* = 5,543 active cases.

The prediction ability of this fitted linear model, the GAM, and the linear and quadratic LOESS models has been evaluated using a 6-fold cross validation procedure, to prevent overfitting. In all cases, the response variable was the estimated number of real COVID-19 active cases in the metropolitan area (Figure 2), and the explanatory variable, the natural logarithm of the viral load. Table 2 shows the corresponding prediction R^2^ for each one of the four models, along with the root mean squared prediction error (RMSPE). The smaller this error, the better the predictive ability of the model was. All the models provided quite accurate predictions for the real number of COVID-19 active cases using the viral load, with an error of around 10% of the response range. The model with the lowest prediction error, 9.5%, was the quadratic LOESS model. Flexible models, such as LOESS and GAM, slightly improved the predictive performance when compared with the linear model, which has a prediction error of around 11.4% of the response range. The quadratic LOESS model was also the one with the largest value for R^2^. Therefore, it provided the best predictive results.

**Table 2.** R^2^ and Root Mean Squared Prediction Error (RMSPE) corresponding to different regression models explaining the number of real active cases in the metropolitan area as a function of the natural logarithm of the viral load. RMSPE was obtained through a 6-fold cross validation procedure.

| Model | $R^2$ | RMSPE |
| --- | --- | --- |
| Linear | 0.8515 | 581.94 |
| GAM | 0.8767 | 508.62 |
| LOESS (linear) | 0.8695 | 487.97 |
| LOESS (quadratic) | 0.8833 | 478.33 |

Figure 11A shows a scatter plot of the estimated number of COVID-19 active cases in the metropolitan area versus the natural logarithm of the viral load, along with the quadratic LOESS fitted curve. Figure 11B displays the actual and predicted values of real number of COVID-19 active cases. The diagonal line was added to compare with the perfect model prediction.

**Figure 11.**
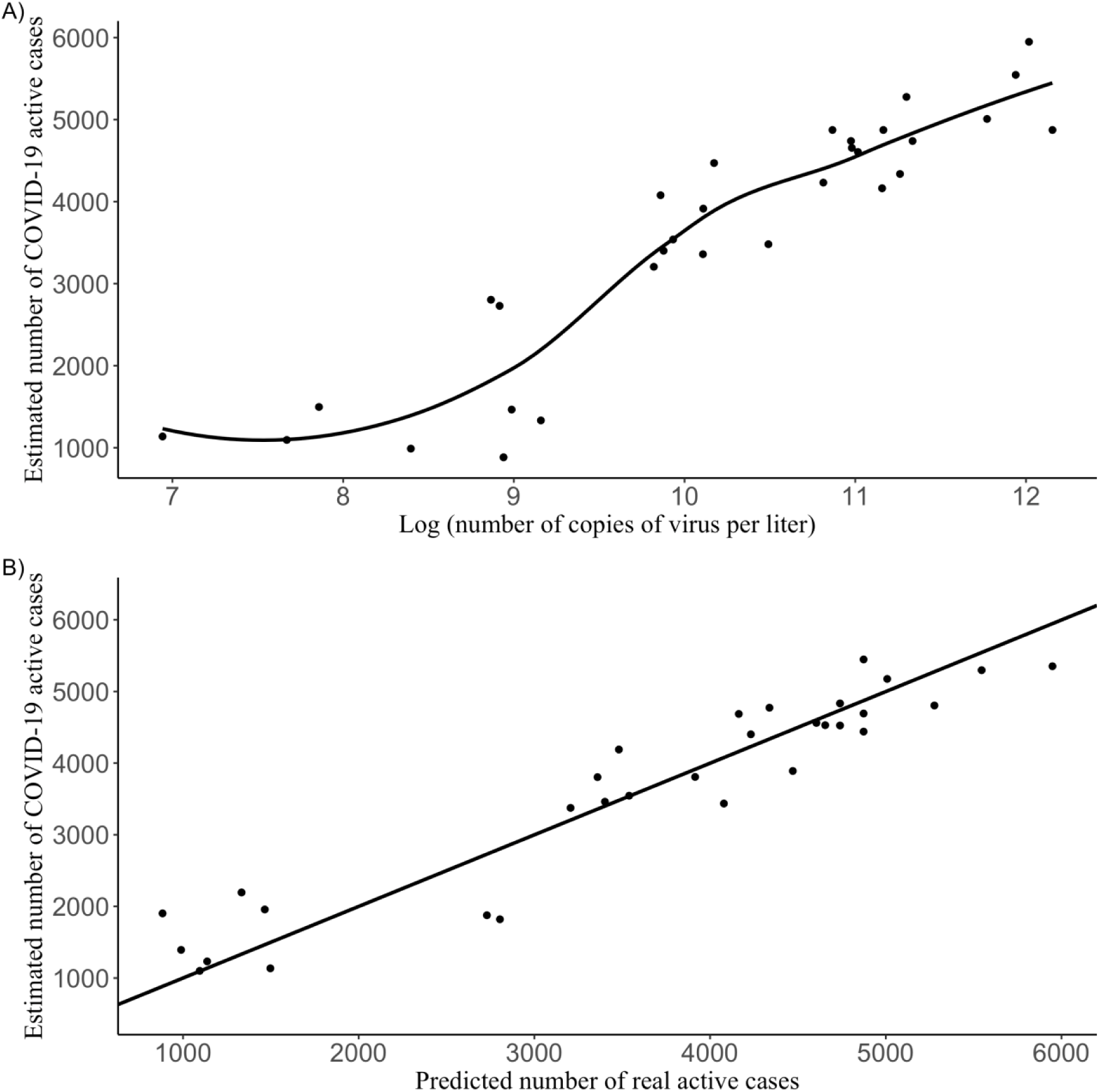
Estimation of number of real COVID-19 active cases using the quadratic LOESS model. A) Estimated COVID-19 active cases in the metropolitan area vs the natural logarithm of the viral load. B) Estimated number of COVID-19 active cases in the metropolitan area vs the predicted number of COVID-19 real active cases.

## 4. DISCUSSION

On March 9^th^ 2020, the city of A Coruña, in the region of Galicia, reported the circulation of SARS-CoV-2 for the first time, with data of a COVID-19 outbreak in a civic center that affected 11 people, as well as data on a few more dispersed cases. At that point, a surveillance phase on approximately 250 people began, and the recommendations from the Health Department on cleaning and mobility restrictions were followed (GCiencia 2020a). During these initial days of the COVID-19 epidemic in Galicia, most of the cases were in the municipality of A Coruña. Thus, at noon on March 13^th^, the Xunta de Galicia (Government of the Autonomous Community of Galicia) reported 90 confirmed cases of COVID-19 in Galicia, 43 of them in the A Coruña area (GCiencia 2020b). The Spanish Government declared a state of alarm on March 14^th^ throughout the country, at which point the Galician community still had few cases. Despite this, in a few days the region went from a monitored and controlled situation to an exponential growth of cases (Rey 2020), reaching a peak of 1667 active cases. The cases were distributed in an area that covers the municipalities of A Coruña and Cee, as shown by the data provided by SERGAS (Galician Health Service) in https://www.datawrapper.de/_/QrkrZ. This area does not coincide with the area that discharges its wastewater into the WWTP Bens, which serves the municipalities of A Coruña, Oleiros, Cambre, Culleredo, and Arteixo, but the figures give an idea of the magnitude of the epidemic at that stage. In this context, an exploratory sampling and analysis was carried out on April 15^th^, which showed the presence of viral genetic material in the wastewater of the WWTP Bens. From April 19^th^, the 24-hour composite samples were continuously analyzed until early June for this study, although surveillance has continued and will continue at WWTP Bens until the virus disappears.

The data from wastewater obtained from April 19^th^ onwards has confirmed the decrease in COVID-19 incidence. We showed that time course quantitative detection of SARS-CoV-2 in wastewater from WWTP Bens correlated with COVID-19 confirmed cases, which backs up the plausibility of our approach. Moreover, the seroprevalence studies carried out by the Spanish Centre for Epidemiology showed that cases in A Coruña represented about 1.8 % of the local population. This means that, for a population of about 369,098 inhabitants, the number of people infected with SARS-CoV-2 contributing their sewage into the WWTP Bens would be around 6,644, which includes people with symptoms and those who are asymptomatic. Considering that the ratio between people with symptoms (reported by the health service) and the total infected population (including asymptomatic people) is estimated to be 1:4, we calculated that reported cases contributing their wastewater into WWTP Bens would be around 1,661, which is close to the maximum number of cases reported in the A Coruña-Cee area (1,667 cases on April 28^th^). It must be noted that the criteria used by the authorities to report cases varied over time, so this may explain the gap between the graphs reported in the media throughout the epidemic and our Figure 6, where both a decrease in the viral load and in the estimated COVID-19 cases can be observed from mid April to early June.

However, the level of the curve at WWTP Bens at the beginning of May was much higher than that corresponding to May 11^th^. This is due to the effectiveness of the lockdown measures applied in Spain. The May 12^th^ daily curve at CHUAC showed a higher viral load than the one corresponding to May 11^th^ at WWTP Bens, showing the viral load measured at the hospital tends to be higher than at WWTP Bens due to a lower dilution effect, as expected.

In the present work, nonparametric and even simple parametric regression models have been shown to be useful tools to construct prediction models for the real number of COVID-19 active cases as a function of the viral load. This is a pioneering approach in the context of the SARS-CoV-2 pandemic since, to our knowledge, WBE studies available are still limited to reporting the occurrence of SARS-CoV-2 RNA in WWTPs and sewer networks, in order to establish a direct comparison with declared COVID-19 cases (Randazzo et al. 2020a, Medema et al. 2020, Nemudryi et al. 2020, La Rosa et al. 2020, Randazzo et al. 2020b, Polo et al. 2020). The only precedent combines computational analysis and modeling with a theoretical approach in order to identify useful variables and confirm the feasibility and cost-effectiveness of WBE as a prediction tool (Hart and Halden 2020). Other examples of WBE models have been applied to previous outbreaks of other infectious diseases. For example, during a polio outbreak detected in Israel in 2013-2014, a disease transmission model was optimized incorporating environmental data (Brouwer et al. 2018). Given the availability of clinical information on poliovirus, the developed infectious disease model incorporated fully validated parameters such as the transmission and vaccination rates, leading to accurate estimations of incidence. This type of study highlights one of the main challenges we have faced developing our model: the SARS-CoV-2 novelty and the associated scarcity of epidemiologic information. Considering this, our statistical model has minimized the uncertainty implementing a complete set of hydraulic information from the sewer network of the city of A Coruña, available thanks of a joint effort from different local authorities. This *ad hoc* model can be adapted to other scenarios as long as similar hydraulic information can be obtained from the area where it will be used.

Other possible explanatory variables (such as rainfall or the mean flow) did not enter the model. Although this is a bit counterintuitive (dilution should affect the viral load measured), it is important to point out that rainfall fluctuated little during the data collection period mid April – early June: its median was 0, its mean was 2.88 L/m^2^ and its standard deviation was 6.59 L/m^2^.

Therefore, as a consequence of the results of the GAM fit, a simple linear model was considered to fit the estimated number of COVID-19 active cases as a function of the logarithm of the viral load. The percentage of variability explained by the model was reasonably high (85.1%) increasing up to 89.4% when three outliers detected were excluded. Alternative, more flexible models, such as GAM and LOESS, were also fitted. They produced slightly better results in terms of R^2^ and RMSPE. The quadratic LOESS model avoided overfitting and showed a good predictive ability (R^2^=0.88, RMSPE=478), the best among all the considered models. However, similar results were found for the linear model that also brings the advantage of simplicity. Therefore, both models, linear and LOESS quadratic, could be successfully used to predict the number of infected people in a given region based on viral load data obtained from wastewater.

Our models, as described, are only applicable to the metropolitan area of A Coruña, the region for which they have been developed, although can be adapted to any other location. This area has Atlantic weather and it may rain substantially in autumn and winter, which could lead to explanatory variables such as rainfall and/or mean flow becoming significant for those seasons and needing to enter the prediction model. Thus, when applying these models to the same location but in seasons with different climatic behavior, they might need to be reformulated. In addition, the methodology used to build these statistical models could be used at other locations for epidemiological COVID-19 outbreak detection, or even for other epidemic outbreaks caused by other microorganisms. Of course, in that case a detailed data analysis would have to be carried out as well, since specific features of the sewage network or the climate may affect the model itself.

## 5. CONCLUSIONS

To the best of our knowledge, these are the first highly reliable wastewater-based epidemiological statistical models useful for tracking COVID-19 epidemic that could be adapted for use anywhere in the world. These models allow the actual number of infected patients to be determined with around 90% reliability, since it takes into account the entire population, whether symptomatic or asymptomatic. These statistical models can estimate the real magnitude of the epidemic at a specific location and their cost-effectiveness and speed of sampling can help to early alert health authorities about potential new outbreaks, thereby helping to protect the local population.

## Data Availability

The authors declare that all data supporting the findings of this study are available within the article and Supplementary Information files, and also are available from the corresponding authors on reasonable request.

## 6. ACKNOWLEDGEMENTS

Authors would like to give special thanks to the Board of Directors from EDAR Bens. Also, we would like to thank Fernanda Rodríguez from the Research Support Services (SAI) at the University of A Coruña, Laura Larriba, from SERGAS, who helped in samples and data collection in CHUAC, Francisco Pérez, Javier Fernández and Cristina Rodríguez from Cadaqua, for their help in sample collection at WWTP Bens, Andrés Paz-Ares and Xurxo Hervada, from SERGAS, who provided the anonymized patient database, Amalia Jácome, Ana López-Cheda, Rebeca Peláez and Wende Safari, from CITIC at UDC, who processed that database to produce the epidemiological series at the municipality level, and Fiona Veira McTiernan, for editing. Finally, we would like to acknowledge the NORMAN European Network for ‘Collaboration in the time of Covid19’.

## FUNDING

This work was supported by EDAR Bens S.A., A Coruña, Spain [grant number INV04020 to MP], the National Plan for Scientific Research, Development and Technological Innovation 2013-2016 funded by the ISCIII, Spain - General Subdirection of Assessment and Promotion of the Research-European Regional Development Fund (FEDER) “A way of making Europe” [grant numbers PI15/00860 to GB and PI17/01482 to MP], the GAIN, Xunta de Galicia, Spain [grant number IN607A 2016/22 to GB, ED431C-2016/015 and ED431C-2020/14 to RC, ED431C 2017/58 to SL, ED431G 2019/01 to RC and SL, and ED431C 2017/66 to MCV], MINECO, Spain [grant number MTM2017-82724-R to RC], the Spanish Network for Research in Infectious Diseases [REIPI RD16/0016/006 to GB]. The work was also supported by the European Virus Archive Global (EVA-GLOBAL) project that has received funding from the European Union’s Horizon 2020 research and innovation program under grant agreement No 871029. SR-F was financially supported by REIPI RD16/0016/006, KC-P by IN607A 2016/22 and the Spanish Association against Cancer (AECC) and JAV by IN607A 2016/22.

## AUTHOR CONTRIBUTIONS

MP, RC, CL, JAV, JT-S and RR conceived and designed the study. JAV, SR-F, MN and KC-P performed wastewater processing and viral analysis, AL-O and JT-S performed statistical models and data analysis, SL managed and analyzed data, BKR-J assisted in the study design and analysis, AA assessed in data collection, AC supervised the wastewater analysis, MCV assessed in wastewater sampling, GB and MP supervised the microbiology team, MP, JAV, RC, RR and JT-S wrote the manuscript. MP and RC supervised the team and coordinated all tasks.

## COMPETING INTERESTS

The authors declare that they have no known competing financial interests or personal relationships that could have appeared to influence the work reported in this paper.

## Notes

### Competing Interest Statement

The authors have declared no competing interest.

### Author Declarations

Authors confirm that it was not necessary to certify the approval of the ethics committee to carry out this work.

## REFERENCES

Ahmed, W., Angel, N., Edson, J., Bibby, K., Bivins, A., O’Brien, J.W., Choi, P.M., Kitajima, M., Simpson, S.L., Li, J., Tscharke, B., Verhagen, R., Smith, W.J.M., Zaugg, J., Dierens, L., Hugenholtz, P., Thomas, K.V. and Mueller, J.F. (2020) First confirmed detection of SARS-CoV-2 in untreated wastewater in Australia: A proof of concept for the wastewater surveillance of COVID-19 in the community. Science of the Total Environ 728, 138764. https://doi.org/10.1016/j.scitotenv.2020.138764

Balboa, S., Mauricio-Iglesias, M., Rodríguez, S., Martínez-Lamas, L., Vasallo, F.J., Regueiro, B. and Lema, J.M. (2020) The fate of SARS-CoV-2 in wastewater treatment plants points out the sludge line as a suitable spot for incidence monitoring. medRxiv, 2020.2005.2025.20112706. https://doi.org/10.1101/2020.05.25.20112706

Bi, Q., Wu, Y., Mei, S., Ye, C., Zou, X., Zhang, Z., Liu, X., Wei, L., Truelove, S.A., Zhang, T., Gao, W., Cheng, C., Tang, X., Wu, X., Wu, Y., Sun, B., Huang, S., Sun, Y., Zhang, J., Ma, T., Lessler, J. and Feng, T. (2020) Epidemiology and transmission of COVID-19 in 391 cases and 1286 of their close contacts in Shenzhen, China: a retrospective cohort study. The Lancet Infectious Diseases 20(8), 911–919. https://doi.org/10.1016/S1473-3099(20)30287-5

Brouwer, A.F., Eisenberg, J.N.S., Pomeroy, C.D., Shulman, L.M., Hindiyeh, M., Manor, Y., Grotto, I., Koopman, J.S. and Eisenberg, M.C. (2018) Epidemiology of the silent polio outbreak in Rahat, Israel, based on modeling of environmental surveillance data. Proceedings of th National Academy of Sciences of the United States of America 115(45), E10625–e10633. https://doi.org/10.1073/pnas.1808798115

Chen, Y., Chen, L., Deng, Q., Zhang, G., Wu, K., Ni, L., Yang, Y., Liu, B., Wang, W., Wei, C., Yang, J., Ye, G. and Cheng, Z. (2020) The presence of SARS-CoV-2 RNA in the feces of COVID-19 patients. Journal of Medical Virology 92(7), 833–840. https://doi.org/10.1002/jmv.25825

Cleveland, W.S. (1979) Robust Locally Weighted Regression and Smoothing Scatterplots. Journal of the American Statistical Association 74(368), 829–836. https://

Croft, T.L., Huffines, R.A., Pathak, M. and Subedi, B. (2020) Prevalence of illicit and prescribed neuropsychiatric drugs in three communities in Kentucky using wastewater-based epidemiology and Monte Carlo simulation for the estimation of associated uncertainties. Journal of Hazardous Materials 384, 121306. https://doi.org/10.1016/j.jhazmat.2019.121306

Day, M. (2020) Covid-19: four fifths of cases are asymptomatic, China figures indicate. The British Medical Journal 369, m1375. https://doi.org/10.1136/bmj.m1375

Ehlers, M.M., Grabow, W.O. and Pavlov, D.N. (2005) Detection of enteroviruses in untreated and treated drinking water supplies in South Africa. Water Research 39(11), 2253–2258. https://doi.org/10.1016/j.watres.2005.04.014

EMCDDA (2020) Perspectives on drugs. Wastewater analysis and drugs: a European multi-city study. https://www.emcdda.europa.eu/topics/pods/waste-water-analysisEnders, C.K. (2010) Applied missing data analysis, Guilford Press, New York, NY, US.

GCiencia (2020a) The outbreak of the civic center of A Coruña accumulates 11 cases of coronavirus (O foco do centro cívico da Coruña suma xa 11 casos de coronavirus). https://www.gciencia.com/saude/centro-civico-coruna-coronavirus/

Gciencia (2020b) Galicia acumulates 90 cases of SARS-CoV-2 (Galicia suma 90 casos de SARS-CoV-2). https://www.gciencia.com/extra/galicia-incidencia-casos-coronavirus/

Goulding, N. and Hickman, M. (2020) A comparison of trends in wastewater-based data and traditional epidemiological indicators of stimulant consumption in three locations. Addiction 115(3), 462–472. https://doi.org/10.1111/add.14852

Gupta, S., Parker, J., Smits, S., Underwood, J. and Dolwani, S. (2020) Persistent viral shedding of SARS-CoV-2 in faeces - a rapid review. Colorectal Disease 22(6), 611–620. https://doi.org/10.1111/codi.15138

Hart, O.E. and Halden, R.U. (2020) Computational analysis of SARS-CoV-2/COVID-19 surveillance by wastewater-based epidemiology locally and globally: Feasibility, economy, opportunities and challenges. Science of the Total Environment 730, 138875. https://doi.org/10.1016/j.scitotenv.2020.138875

Hastie, T. and Tibshirani, R. (1990) Generalized Additive Models, Chapman and Hall. https://doi.org/10.1002/sim.4780110717

Hellmér, M., Paxéus, N., Magnius, L., Enache, L., Arnholm, B., Johansson, A., Bergström, T. and Norder, H. (2014) Detection of pathogenic viruses in sewage provided early warnings of hepatitis A virus and norovirus outbreaks. Applied and Environmental Microbiology 80(21), 6771–6781. https://10.1128/AEM.01981-14

Hovi, T., Shulman, L.M., van der Avoort, H., Deshpande, J., Roivainen, M. and EM, D.E.G. (2012) Role of environmental poliovirus surveillance in global polio eradication and beyond. Epidemiology & Infection 140(1), 1–13. https://10.1017/S095026881000316X

La Rosa, G., Iaconelli, M., Mancini, P., Bonanno Ferraro, G., Veneri, C., Bonadonna, L., Lucentini, L. and Suffredini, E. (2020) First detection of SARS-CoV-2 in untreated wastewaters in Italy. Science of Total Environment 736, 139652. https://doi.org/10.1016/j.scitotenv.2020.139652

Lizasoain, A., Tort, L.F.L., García, M., Gillman, L., Alberti, A., Leite, J.P.G., Miagostovich, M.P., Pou, S.A., Cagiao, A., Razsap, A., Huertas, J., Berois, M., Victoria, M. and Colina, R. (2018) Human enteric viruses in a wastewater treatment plant: evaluation of activated sludge combined with UV disinfection process reveals different removal performances for viruses with different features. Letters in Applied Microbiology 66(3), 215–221. https://doi.org/10.1111/lam.12839

Lodder, W. and de Roda Husman, A.M. (2020) SARS-CoV-2 in wastewater: potential health risk, but also data source. The Lancet Gastroenterology Hepatology 5(6), 533–534. https://doi.org/10.1016/S2468-1253(20)30087-X

Mancini, P., Bonanno Ferraro, G., Iaconelli, M. and Suffredini, E. (2019) Molecular characterization of human Sapovirus in untreated sewage in Italy by amplicon-based Sanger and next-generation sequencing. Journal of Applied Microbiology 126(1), 324–331. https://doi.org/10.1111/jam.14129

Medema, G., Heijnen, L., Elsinga, G., Italiaander, R. and Brouwer, A. (2020) Presence of SARS-Coronavirus-2 RNA in Sewage and Correlation with Reported COVID-19 Prevalence in the Early Stage of the Epidemic in The Netherlands. Environmental Science & Technology Letters 7(7), 511–516. https://doi.org/10.1021/acs.estlett.0c00357

Nemudryi, A., Nemudraia, A., Wiegand, T., Surya, K., Buyukyoruk, M., Cicha, C., Vanderwood, K.K., Wilkinson, R. and Wiedenheft, B. (2020) Temporal Detection and Phylogenetic Assessment of SARS-CoV-2 in Municipal Wastewater. Cell Reports Medicine 1(6), 100098. https://doi.org/10.1016/j.xcrm.2020.100098

Peccia, J., Zulli, A., Brackney, D.E., Grubaugh, N.D., Kaplan, E.H., Casanovas-Massana, A., Ko, A.I., Malik, A.A., Wang, D., Wang, M., Warren, J.L., Weinberger, D.M., Arnold, W. and Omer, S.B. (2020) Measurement of SARS-CoV-2 RNA in wastewater tracks community infection dynamics. Nature Biotechnology 38(10), 1164–1167. https://10.1038/s41587-020-0684-z

Pollán, M., Pérez-Gómez, B., Pastor-Barriuso, R., Oteo, J., Hernán, M.A., Pérez-Olmeda, M., Sanmartín, J.L., Fernández-García, A., Cruz, I., Fernández de Larrea, N., Molina, M., Rodríguez-Cabrera, F., Martín, M., Merino-Amador, P., León Paniagua, J., Muñoz-Montalvo, J.F., Blanco, F. and Yotti, R. (2020) Prevalence of SARS-CoV-2 in Spain (ENE-COVID): a nationwide, population-based seroepidemiological study. The Lancet 396(10250), 535–544. https://doi.org/10.1016/S0140-6736(20)31483-5

Polo, D., Quintela-Baluja, M., Corbishley, A., Jones, D.L., Singer, A.C., Graham, D.W. and Romalde, J.L. (2020) Making waves: Wastewater-based epidemiology for COVID-19 - approaches and challenges for surveillance and prediction. Water Research 186, 116404. https://doi.org/10.1016/j.watres.2020.116404

R Core Team: A language and environment for statistical computing, R Foundation for Statistical Computing, Vienna, Austria. https://www.r-project.org/

Randazzo, W., Truchado, P., Cuevas-Ferrando, E., Simón, P., Allende, A. and Sánchez, G. (2020a) SARS-CoV-2 RNA in wastewater anticipated COVID-19 occurrence in a low prevalence area. Water Research 181, 115942. https://doi.org/10.1016/j.watres.2020.115942

Randazzo, W., Cuevas-Ferrando, E., Sanjuán, R., Domingo-Calap, P. and Sánchez, G. (2020b) Metropolitan wastewater analysis for COVID-19 epidemiological surveillance. International Journal of Hygiene Environmental Health 230, 113621. https://doi.org/10.1016/j.ijheh.2020.113621

Rey, M. (2020) María José Pereira: “We are ready, but citizen responsability is essential” GCiencia https://www.gciencia.com/saude/maria-jose-pereira-estamos-preparados-pero-a-responsabilidade-cidada-sera-esencial/

Schloerke, B., Crowley, J., Cook, D., Hofmann, H., Wickham, H., Briatte, F., Marbach, M., Thoen, E., Elberg, A., Larmarange, J. and Toomet, O. (2020) GGally: Extension to ‘ggplot2’. https://ggobi.github.io/ggally/

Wickham, H. (2016) ggplot2: Elegant Graphics for Data Analysis, Springer International Publishing.

Wölfel, R., Corman, V.M., Guggemos, W., Seilmaier, M., Zange, S., Müller, M.A., Niemeyer, D., Jones, T.C., Vollmar, P., Rothe, C., Hoelscher, M., Bleicker, T., Brünink, S., Schneider, J., Ehmann, R., Zwirglmaier, K., Drosten, C. and Wendtner, C. (2020) Virological assessment of hospitalized patients with COVID-2019. Nature 581(7809), 465–469. https://10.1038/s41586-020-2196-x

Wood, S. (2006) Generalized Additive Models: An Introduction with R, Taylor & Francis. https://doi.org/10.1201/9781315370279

Wu, Y., Guo, C., Tang, L., Hong, Z., Zhou, J., Dong, X., Yin, H., Xiao, Q., Tang, Y., Qu, X., Kuang, L., Fang, X., Mishra, N., Lu, J., Shan, H., Jiang, G. and Huang, X. (2020a) Prolonged presence of SARS-CoV-2 viral RNA in faecal samples. The Lancet Gastroenterology & Hepatology 5(5), 434–435. https://doi.org/10.1016/S2468-1253(20)30083-2

Wu, F., Zhang, J., Xiao, A., Gu, X., Lee, W.L., Armas, F., Kauffman, K., Hanage, W., Matus, M., Ghaeli, N., Endo, N., Duvallet, C., Poyet, M., Moniz, K., Washburne, A.D., Erickson, T.B., Chai, P.R., Thompson, J. and Alm, E.J. (2020b) SARS-CoV-2 Titers in Wastewater Are Higher than Expected from Clinically Confirmed Cases. mSystems 5(4). https://10.1128/mSystems.00614-20

Wurtzer, S., Marechal, V., Mouchel, J.-M., Maday, Y., Teyssou, R., Richard, E., Almayrac, J.L. and Moulin, L. (2020) Evaluation of lockdown impact on SARS-CoV-2 dynamics through viral genome quantification in Paris wastewaters. medRxiv, 2020.2004.2012.20062679. https://doi.org/10.1101/2020.04.12.20062679

Xing, Y.H., Ni, W., Wu, Q., Li, W.J., Li, G.J., Wang, W.D., Tong, J.N., Song, X.F., Wing-Kin Wong, G. and Xing, Q.S. (2020) Prolonged viral shedding in feces of pediatric patients with coronavirus disease 2019. Journal of Microbiology, Immunology and Infection 53(3), 473–480. https://doi.org/10.1016/j.jmii.2020.03.021

Xu, Y., Li, X., Zhu, B., Liang, H., Fang, C., Gong, Y., Guo, Q., Sun, X., Zhao, D., Shen, J. and Zhang, H. (2020) Characteristics of pediatric SARS-CoV-2 infection and potential evidence for persistent fecal viral shedding. Nature Medicine 26(4), 502–505. https://10.1038/s41591-020-0817-4

Yang, R., Gui, X. and Xiong, Y. (2020) Comparison of Clinical Characteristics of Patients with Asymptomatic vs Symptomatic Coronavirus Disease 2019 in Wuhan, China. Journal of American Medical Association Network Open 3(5), e2010182. https://10.1001/jamanetworkopen.2020.10182

Zhang, T., Cui, X., Zhao, X., Wang, J., Zheng, J., Zheng, G., Guo, W., Cai, C. and He, S. (2020) Detectable SARS-CoV-2 viral RNA in feces of three children during recovery period of COVID-19 pneumonia. Journal of Medical Virology 92(7), 909–914. https://doi.org/10.1002/jmv.25795

